# Caregiver’s Decision to Report Adverse Drug Reactions among Children Receiving Seasonal Malaria Chemoprevention in Ghana

**DOI:** 10.1101/2024.11.28.24318172

**Authors:** Abdul Gafaru Mohammed, Dora Dadzie, George Adu Asumah, Isaac Adomako, Joel Jeffrey Idun-Acquah, Paul Boateng, Nana Yaw Peprah, Keziah L. Malm

## Abstract

**Introduction:** The antimalarial medicines used in Seasonal Malaria Chemoprevention (SMC) campaigns are generally well-tolerated but adverse drug reactions (ADRs) can occur. Monitoring, reporting, and prompt management of ADRs is essential to build and maintain trust in SMC campaigns in the implementing communities. The caregiver’s decision to report ADRs represents a critical issue at the intersection of public health, child welfare, and pharmacovigilance. As SMC campaigns continue to expand, it is imperative to understand and address the factors influencing caregivers’ decisions to report ADRs.

**Methods:** A mixed methods cross-sectional study involving questionnaire administration, focus group discussions, and review of children’s health records was employed to collect data from caregivers of children aged 3-59 months in the Northern, North-East, and Savanna regions of Ghana. A systematic random sample of 679 caregivers was recruited for the study across three regions. Data including ADR occurrence, maternal and child characteristics was collected from selected caregivers in their homes. Logistic regression was performed for associations between caregiver’s reports of ADRs and caregiver characteristics.

**Results:** About 49.5% (336/679) of caregivers mentioned the occurrence of ADRs in children after receiving SMC medication. The commonly cited ADR was diarrhea (34.7%, 116/336). Only 16.9% (57/336) of caregivers reported ADRs to the health workers at the time of occurrence. After adjusting for confounders, increasing age of child (aOR=1.04, 95%CI:1.008-1.065), receiving education on ADR reporting (aOR=4.03, 95%CI:4.366-6.119), education on management of mild ADRs (aOR=4.43, 95%CI:2.094-9.808) and having a means of reaching health personnel (aOR=1.56, 95%CI:1.202-2.037) increased the odds of ADR reporting while increasing age of caregivers (aOR=0.92, 95%CI:0.883-0.966) and being married (aOR=0.17, 95%CI:0.149-0.183) decreased the odds of reporting ADRs among the caregivers studied.

**Conclusion:** Less than 20% of caregivers whose children experienced ADRs after receiving SMC medication reported the incident. Caregiver education on ADRs and it’s management and means of reaching the health team were modifiable factors that influenced caregiver ADR reporting.

## Introduction

To complement ongoing malaria control efforts, including vector control measures, prompt diagnosis of suspected malaria, and treatment of confirmed cases with antimalarial medicines, the WHO in 2012 recommended seasonal malaria chemoprevention (SMC) for the control of malaria among children under age 5 (1–3) in areas where malaria transmission is seasonal. SMC is defined as “the intermittent administration of full treatment of an anti-malarial medicine during the malaria season to prevent malarial illness and to maintain therapeutic drug concentrations in the blood throughout the greatest malarial risk (4). Ghana adopted this preventive intervention in 2013 (2) for children aged 3-59 months. SMC began with a pilot from July 2015 to October 2015 in the Lawra district of the Upper West region, followed by a full-scale implementation in the year 2016 across all districts of the Upper West and Upper East regions (2). Currently, the intervention has been implemented in 69 districts located in Northern, North-East, Savannah, Upper East, Upper West, Bono East, and Oti regions of Ghana.

The success of SMC depends on the efficacy and safety of the amodiaquine plus sulfadoxine-pyrimethamine (AQ+SP) (5,6). Countries implementing SMC have adopted various pharmacovigilance reporting measures either passive or active for reporting possible adverse drug reactions (7,8). Ghana adopted a passive pharmacovigilance reporting process for SMC-related ADRs whose success largely depends on the caregivers of the children receiving the therapy.

SMC administration has been reported to be associated with various adverse reactions among children receiving the therapy (9–11). The most commonly reported side effects are diarrhea, abdominal pain, body weakness and vomiting (8). These ADRs have been reported by caregivers to mostly occur within the second or third day after administering the therapy. ADR reporting rate has been reported to be low among health workers and caregivers of children receiving the therapy (12–14). In a study conducted in northern Nigeria, out of seven states implementing SMC, only two states reported ADRs from the exercise (11).

Reporting adverse drug reactions by caregivers of children receiving SMC has been reported to be influenced by various factors. These factors include poor knowledge, not being aware of the term ADR, single marital status, lower education level, and difficulty accessing public clinics for medical services (15).

There is currently a paucity of data on the reporting of ADRs among caregivers of children receiving SMC and the factors that influence their decision to report. Studies conducted on ADRs among children receiving SMC in the West African sub-region focus on the healthcare staff providing the service with none exploring the problem among caregivers (11). With the passive pharmacovigilance reporting approach adopted for SMC ADRs in Ghana, it is important to assess the rate of ADR reporting among caregivers of children under the therapy and also the factors that influence their decision to report ADRs. This study was therefore conducted to assess the factors that influence the decision of caregivers to report ADRs when they occur among children receiving SMC in Ghana.

## Methods

### Study design

This was a mixed methods cross-sectional study among caregivers of children between 6 – 59 months in Ghana. A semi-structured questionnaire, data abstraction tool and FGD guide were used to collect data on the characteristics of children and their caregivers, the uptake of SMC, the occurrences of ADRs, and its reporting.

### Study setting

The study was conducted in Ghana. Ghana is located in West Africa with an estimated 31 million people, 16 administrative regions, and 261 districts. The country shares boundaries with Togo, Burkina Faso, Ivory Coast, and the Gulf of Guinea. Following the WHO recommendation of SMC as an intervention, Ghana adopted it in 2013 and included it in the NSP for implementation to ensure that malaria morbidity and mortality among children under five years are reduced to zero. Malaria cases among children under five have seen a drastic decline in the implementing regions since the intervention was rolled out. The intervention is usually given during the transmission period (rainy season). In 2021 and 2022, Ghana Health Service (GHS) through the National Malaria Elimination Programme supported 7 regions (Northern, North-East, Savannah, Upper East, Upper West, Bono East, and Oti) to implement the SMC in 72 districts. The study was conducted in three of the seven regions. These regions included Savanna, Northern, and North East Regions. In 2022 and 2023, each of these regions received four cycles of seasonal malaria chemoprevention within the rainy season. The rainy season ranged from May – September.

### Study population

The study included all caregivers of children aged 3 – 59 months in the Northern, Savanna, and North East Region. in Ghana. Caregivers of children between 3 – 59 months of age, who have lived in the selected districts for at least one year were included in the study. Caregivers of children aged 3 – 59 months in whom the use of SMC is contraindicated were excluded from the study. Also, caregivers of children who missed the 2023 SMC dosing campaign were excluded from the study.

### Sample size estimation

Using Z = indicator for a confidence level (1.96 for 95% confidence level), n = minimum sample size, De is the design effect, the ratio between the variance from the cluster design to the variance that would be obtained from a simple random sampling, Z (1.96), P (66.5%) (16) and e (5%) is the margin of error allowed. We estimated a minimum sample size using the formula n = [De × Z^2^ × p (1-p)]/e^2^. n = [2 × (1.96)^2^ × 0.67× 0.33]/(0.05)^2^ = 679.

### Sampling process

A multistage cluster sampling approach was used to sample caregivers for the study. The first stage involved the simple random sampling of three (3) regions out of the 7 regions implementing SMC. Followed by sampling of 4 districts from each of the three (3) selected regions. A stratified random sampling approach was then used to sample two communities from each of the districts. Two strata were formed for each district, stratum A (list of rural communities in the district) and stratum B (list of urban communities in the district). One community was randomly selected from each of these strata making up two communities from each district. A probability proportionate to size sampling approach using the number of children registered for the 2022 SMC in each of the selected communities was used to determine the total number of caregivers to interview in each of the communities. Finally, a systematic random sampling approach was used to recruit caregivers for interviews on each day of data collection by trained research assistants.

### Data collection

Data was collected from caregivers of children between 3 – 59 months using a pre-tested semi-structured questionnaire, a focus group discussion guide, and a data abstraction tool adapted from previous studies (17–19). Data was collected between October – December 2023. The questionnaire was prepared and deployed on Kobo collect for data collection. The questionnaire was prepared in English and interpreted into Dagare, Kasena, Frafra, Twi, Asante, Dagbani, Konkomba, and Nanumba during administration. The semi-structured questionnaire elicited data on children’s characteristics (age, sex, education), characteristics of their caregivers (age, sex, marital status, region, residence, educational status, employment status), awareness and knowledge of ADRs, and occurrence of ADRs and its reporting. The data abstraction tool was used to extract data on the update of SMC and the number of doses received from the maternal and child health records book.

Fifteen community health nurses were recruited and trained as research assistants to collect data from the study participants. The research assistants were trained in the general overview of the study, the sampling approach, the eligibility procedure, all sections of the questionnaire, questionnaire administration, the use of data abstraction tool, participant privacy and confidentiality, and their safety. The data collection tools were pre-tested among 30 caregivers in Ahafo Region prior to the actual data collection. All issues observed during the pre-test were rectified before the actual data collection.

### Data analysis

The data collected was extracted from Kobo-Collect as Microsoft Excel files, cleaned, and imported into R-studio for statistical analysis. The data was descriptively analyzed, and categorical variables were summarized into frequencies and percentages with accompanying 95% Confidence Intervals. Parametric continuous variables such as age were summarized into means and standard deviations. A crude binary logistic regression analysis was performed to determine the association between participants’ characteristics and the decision to report ADRs. Variables with p< 0.25 were considered statistically significant and selected for an adjusted binary logistic regression analysis. At the adjusted level, the level of significance was set at 5%.

## Results

### Background characteristics of caregivers and children

Out of the 679 caregivers studied, majority (57.6%; 391) were rural dwellers. The median age of caregivers was 29 (IQR= 25 – 33) years with most (63.5%; 431) being between 25 – 34 years old. Almost half (45.8%; 311) of the caregivers were housewives or unemployed. The average age of the children under the care of selected caregivers was 28 (IQR=20 – 34) months with majority (52.4%; 356) of them being male (**Table 1**).

**Table 1:** Background characteristics of caregivers and children.

| Characteristics | Frequency (n = 679) | Proportion (%) |
| --- | --- | --- |
| **Place of residence** |  |  |
| Rural | 391 | 57.58 |
| Urban | 288 | 42.42 |
| **Age of caregiver, median (IQR) years** | 29 (25 – 33) |  |
| 18 – 24 | 132 | 19.44 |
| 25 – 34 | 431 | 63.48 |
| 35+ | 116 | 17.08 |
| **Educational level** |  |  |
| No formal education | 256 | 37.70 |
| Primary | 166 | 24.45 |
| Secondary | 172 | 25.33 |
| Tertiary | 85 | 12.52 |
| **Occupation** |  |  |
| Government employed | 100 | 14.73 |
| Unemployed or housewife | 311 | 45.80 |
| Trader | 268 | 39.47 |
| **Marital status** |  |  |
| Married | 617 | 90.87 |
| Unmarried | 62 | 9.13 |
| **Age of child [median (IQR) months]** | 28 (20 – 34) |  |
| **Sex of child** |  |  |
| Female | 323 | 47.57 |
| Male | 356 | 52.43 |

### Individual-level characteristics of the caregivers studied

More than two-thirds (70.1%; 476) of the caregivers had a health facility in their community of residence. Also, almost half (47.9%; 325) of the caregivers had no means of contacting the medical team in case of a severe adverse event (**Table 2**).

**Table 2:** Individual-level characteristics of the caregivers studied.

| Characteristics | Frequency (n = 679) | Proportion (%) |
| --- | --- | --- |
| **Availability of HF** |  |  |
| No | 203 | 29.90 |
| Yes | 476 | 70.10 |
| **Received education on ADR reporting** |  |  |
| No | 327 | 48.16 |
| Yes | 352 | 51.84 |
| **Received education on management of mild ADRs** |  |  |
| No | 262 | 38.59 |
| Yes | 417 | 61.41 |
| **Means of contacting health workers** |  |  |
| No | 325 | 47.86 |
| Yes | 354 | 52.14 |

### Occurrence of Adverse drug reactions and reporting

Out of 679 caregivers studied, almost half (49.5%; 336) mentioned the occurrence of adverse drug reactions in their children following the uptake of SMC medications in the most recent SMC cycle. The most common ADRs experienced were fever (44.2%; 148) and diarrhea (34.6%; 116). Majority (48.4%; 162) of these ADRs occurred after 2 hours of receiving medications with (41.1%; 138) managing their children at home. Less than a fifth (16.9%; 57) of the caregivers reported the ADRs to the nearest health facility or a medical practitioner (**Table 3**).

**Table 3:** Occurrence of Adverse drug reactions and reporting.

| Variable | Frequency (n = 679) | Proportion (%) |
| --- | --- | --- |
| **Adverse drug reaction (n = 679)** |  |  |
| Did not occur | 343 | 50.52 |
| Occurred | 336 | 49.48 |
| **Type of ADR (n = 336)** |  |  |
| Diarrhea | 116 | 34.63 |
| Vomiting | 96 | 28.57 |
| Headache | 28 | 8.33 |
| Rashes | 22 | 6.55 |
| Fever | 74 | 22.02 |
| **Time of occurrence (n = 335)** |  |  |
| Within 1 | 84 | 25.07 |
| 1 – 2 hours | 89 | 26.57 |
| After 2 hours | 162 | 48.36 |
| **Action taken (n = 336)** |  |  |
| Visited drug store | 93 | 27.68 |
| Managed child at home | 138 | 41.07 |
| Did nothing | 73 | 21.73 |
| Take child to health facility | 32 | 9.52 |
| **Reporting (n = 336)** |  |  |
| No | 279 | 83.04 |
| Yes | 57 | 16.96 |

### Stratification of ADR Reporting by place of residence

Overall, out of 336 children who experienced ADRs after receiving SMC medication, only 16.9 (95%CI: 0.132 – 0.214) reported the occurrence. Reporting of ADRs was higher among caregivers residing in urban settings compared to their counterparts in rural settings (**Table 4**).

**Table 4:** Stratification of ADR Reporting by place of residence.

| Residence | Proportion of ADR reporting | 95% CI |
| --- | --- | --- |
| Rural | 1.6 | 0.003 0.046 |
| Urban | 36.0 | 0.283 0.442 |
| Overall | 16.9 | 0.132 0.214 |

### Factors associated with associated with ADRs Reporting among caregivers

After adjusting for the effect of confounders, age of caregivers (aOR = 0.92, 95%CI: 0.883 - 0.966) marital status (aOR = 0.17, 95%CI: 0.149 - 0.183), age of child (aOR = 1.04, 95%CI: 1.008 - 1.065), education on ADR reporting (aOR = 4.03, 95%CI: 4.366 - 6.119) and management of mild ADRs (aOR = 4.43, 95%CI: 2.094 - 9.808), and means of reaching health personnel (aOR = 1.56, 95%CI: 1.202 - 2.037) were factors found to be significantly associated with the reporting of ADRs among the caregivers studied. Caregivers who were educated on ADR reporting had 4 times increased odds of reporting adverse events when they occurred compared to those who were not educated on ADR reporting (aOR = 4.03, 95%CI: 4.366 - 6.119). Also, A unit increase in the age of caregivers was associated with an 8% decrease in the odds of reporting adverse events following SMC in their children (aOR = 0.92, 95%CI: 0.883 - 0.966) (**Table 5**).

**Table 5:**
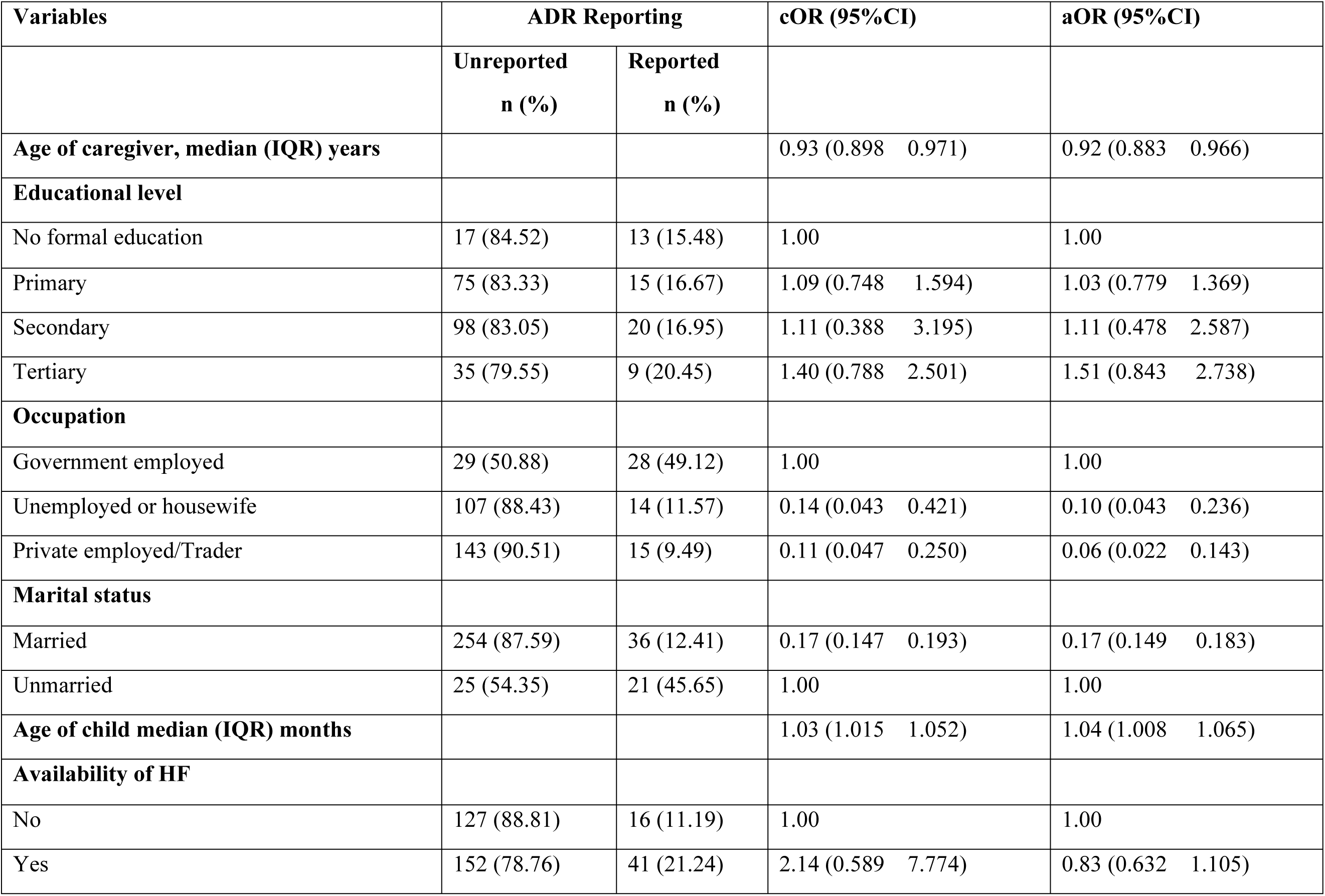
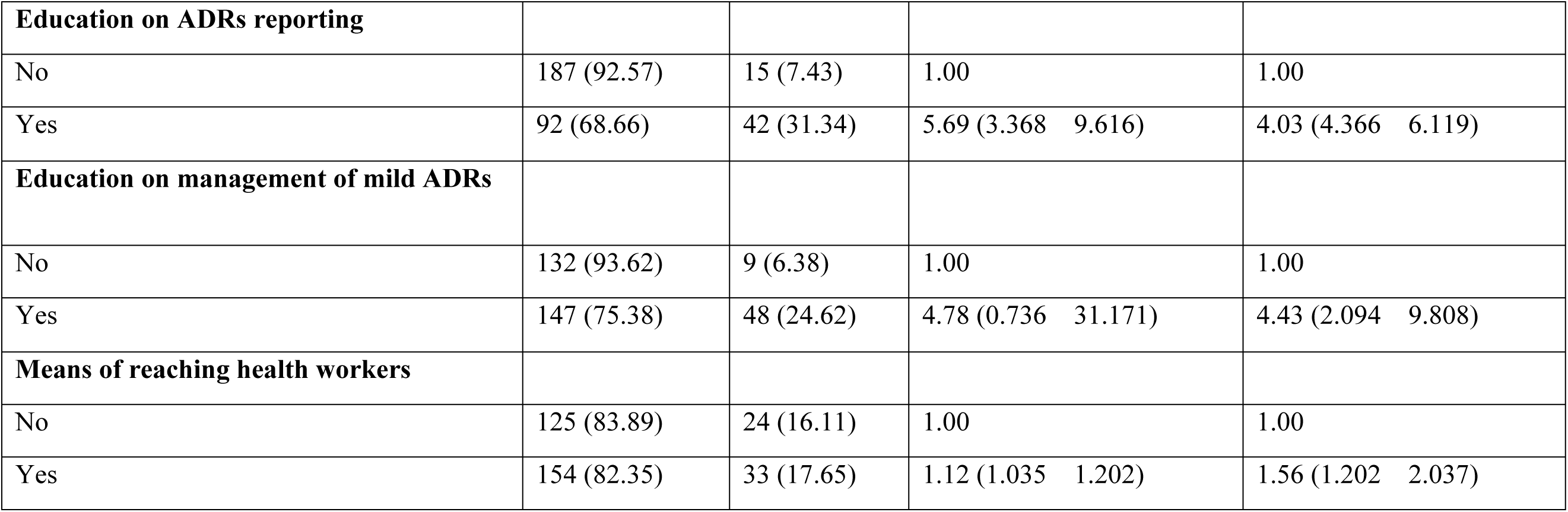
Factors associated with associated with ADRs Reporting among Caregivers.

### Occurrence of ADRs and its reportage among caregivers

*“The health worker told us the medicines will protect our children against malaria. I give the drugs to both my children; I wasn’t informed my children may have any reactions after taking the medicines”*. **Caregiver 4**

*“I noticed that my child’s appetite decreased after taking the medication on the first day. He even refused his favorite food which I prepared for him. I thought this was because he had started growing teeth, I didn’t know the drugs could affect his appetite I would have reported to the health facility if I knew”*. **Caregiver 6**

*“My child started running diarrhea, the same day the health workers came to the house to give him the drugs, the diarrhea increased after I gave him the other one on the second day so I didn’t even give him the last one, I was scared”*. **Caregiver 1**

*“The man who came to our house to give the medicine to my son told me he might vomit or have a hot temperature or run diarrhea so I should go to the hospital if it happened. Nothing happened on the first day but my son had a hot temperature the third day after I gave him the drugs. I gave him paracetamol syrup I got the last time he was sick”*. **Caregiver 5**

*“My husband is a nurse, so when my daughter had diarrhea after taking the drugs on the second day, I informed him, and he said maybe the diarrhea is caused by the drug so I should take her to the health center in our area. I took her there and the health workers gave us some drugs, they didn’t even charge us money that day”*. **Caregiver 10**

## Discussion

Seasonal administration of antimalaria drug, sulphadoxine/pyrimethamine plus amodiaquine (SPAQ) to children 3–59 months is a tool in the reduction of childhood malaria morbidity and mortality in areas with highly seasonal malaria transmission like Northern Ghana (20). In 2023, 1,492,779 children in the Northern, North East, Upper West, Upper East, Bono East, and Oti regions received SPAQ during the fourth round of the seasonal malaria chemoprevention campaign in Ghana. Using a mixed methods approach, our study assessed the factors that influence the decision of caregivers to report ADRs when they occur among children receiving SMC in Ghana. Less than 17% of caregivers were found to report adverse drug reactions when they occurred in this study. The reporting rate found in this study is consistent with the findings of similar studies conducted in Northern Nigeria and Senegal where ADR reporting was found to be abysmally low (21,22). Also, in a study assessing parental reporting of adverse drug reactions in South Africa, majority (66.5%) of participants did report an ADR to a healthcare professional (23).

The occurrence of ADRs during SMC that go unreported or unresolved tends to affect the trust caregivers have on SMC medications thereby affecting the willingness of caregivers to allow their children to receive SMC medications. The ready availability of health workers or volunteers to caregivers tends to increase ADR reporting. To achieve this, the National Malaria Elimination Programme through the various regional and district health directorates needs to implement and strengthen follow-up visits to the homes of children receiving SPAQ during SMC campaigns.

The commonly reported ADRs were fever, diarrhea, and vomiting. This is corroborated by the findings of a similar study among young children in Northern Sahelian Ghana where diarrhea, vomiting, and fever were the most experienced adverse events during the four rounds of SMC administration (24). Also, in Community-Based Safety Monitoring during Seasonal Malaria Chemoprevention Campaigns in Senegal, the most commonly reported symptoms were vomiting, fever, and abdominal pain (21). These reported ADRs have the tendency of putting a child in a dire situation if not managed properly and timely. During SMC campaigns, caregivers need to be educated on the reporting of these adverse events and also first aid management of these events prior to the arrival of the medical team.

Our study found a significant association between marital status and ADR reporting among the caregivers studied. Caregivers who were unmarried had increased odds of reporting ADRs compared to married caregivers. The decision-making power in Northern Ghana mostly lies with husbands in marriage. Caregivers who are married mostly need to seek advice from their partners before taking any health-seeking decision, this may delay or possibly impede the reporting of adverse events. Unlike married caregivers, unmarried caregivers mostly have the decision-making power lying with them and they can easily report ADRs when they occur without consulting another person.

The ease of access to health personnel and healthcare facilities can influence parental reporting of ADRs. Caregivers who have convenient means of reaching healthcare providers, such as through telephone hotlines, online portals, or direct visits to healthcare facilities, may be more likely to report adverse events. Accessibility to health personnel can facilitate timely reporting of adverse events and increase the likelihood of receiving appropriate guidance on managing these events. Our study found increased odds of reporting adverse events among caregivers with means of reaching the health team compared to caregivers without any means of reaching the health authority. To increase ADR reporting, district health directorates need to adopt a strategy where volunteers share the call lines of health facilities located in the community during SMC campaigns in those communities. This will facilitate ease of reporting ADRs when they occur.

Furthermore, education on ADR reporting was also found to increase the reporting of ADRs among caregivers. This is consistent with the findings of a study among pharmacists in Egypt where those who received education on adverse events reporting had an increased chance of reporting events compared to those who were not educated on the reporting process (19). Similarly, education on adverse events reporting was reported to increase adverse event identification and reporting among nursing health professionals in a tertiary health facility in India (25). This implies that increasing caregiver awareness of adverse events and its reporting process will increase the number of caregivers who would report adverse events when they occur in their children during SMC. Also, education on the management of mild adverse events was another factor found to be significantly associated with adverse events reporting by caregivers in this study. Caregivers who received education on how to manage mild or minor adverse events had increased odds of reporting adverse events compared to those not educated. Health volunteers visiting homes during SMC should be strengthened to provide education on ADR, management, and reporting during dosing.

A limitation of this study was recall bias, caregivers recalled ADRs in their children who received medication in the past, and this could have led to underestimation or overestimation of ADR occurrence and its reporting.

## Conclusion

Less than 20% of caregivers whose children experienced ADRs after receiving SMC medication reported the incident. Caregiver education on ADRs and it’s management and means of reaching the health team were modifiable factors that influenced the caregiver’s ADR reporting.

## Data Availability

All relevant data are within the manuscript and its Supporting Information files.

## Acknowledgments

We wish to acknowledge the research assistants and the various regional and district health directorates that supported the exercise.

## Authors’ contributions

AGM & DD conceptualized the study and conducted data. AGM and PB analyzed the collected data. GAA, IA, JJIA, NYP and KLM, drafted the initial manuscript. All authors read and approved the manuscript for publication.

## Availability of data and materials

The data for this study is available in this publication.

## Declarations

Ethical clearance for the study was obtained from the Ghana Health Service Ethical Review Committee. Permission was sought from the various regional and district health directorates in the region before the study. Written informed consent was obtained from the caregivers of children recruited for the study. The data collected was devoid of personal identifiers and was used solely for this study.

## Consent for publication

Not applicable.

